# Glymphatic System in Temporal Lobe Epilepsy Associated with Encephalocele

**DOI:** 10.64898/2026.07.02.26356654

**Authors:** Roberta Di Giacomo, Dalila Biancheri, Alessandra Burini, Fabio Martino Doniselli, Laura Rossini, Elisa Visani, Valeria Cuccarini, Gianluca Marucci, Annalisa Parente, Giuseppe Didato, Francesco Deleo, Chiara Pastori, Giulia Battaglia, Giulia Maccanti, Giulia Sofia Cereda, Michele Rizzi, Marco de Curtis, Rita Garbelli

**Affiliations:** Epilepsy Unit, Fondazione IRCCS Istituto Neurologico Carlo Besta, Milan, Italy; Neurology Unit, Department of Medicine (DMED), University of Udine, Udine, Italy; Neuroradiology Unit, Fondazione IRCCS Istituto Neurologico Carlo Besta, Milan, Italy; Neuropathology Unit, Fondazione IRCCS Istituto Neurologico Carlo Besta, Milan, Italy; Neurosurgery Unit, Fondazione IRCCS Istituto Neurologico Carlo Besta, Milan, Italy

**Author notes:** These authors equally contributed to the present work (Roberta Di Giacomo, Dalila Biancheri, Alessandra Burini, Fabio Martino Doniselli). **Correspondence to**: Fabio Martino Doniselli, Neuroradiology Unit, Fondazione IRCCS Istituto Neurologico Carlo Besta, Dalila Biancheri, Epilepsy Unit, Fondazione IRCCS Istituto Neurologico Carlo Besta.

**Keywords:** Temporal lobe epilepsy, encechephalocele, glymphatyc system

## Abstract

**Objective:** Temporal lobe encephaloceles (ENC) are underdiagnosed causes of drug-resistant temporal lobe epilepsy (TLE), frequently associated with idiopathic intracranial hypertension (IIH). Emerging evidence suggests glymphatic system dysfunction in both IIH and TLE. We investigated glymphatic markers in TLE associated with ENC compared with seizure-free postoperative TLE controls of different aetiology.

**Methods:** Surgical specimens from 13 patients with TLE-ENC and 12 TLE-control patients were analyzed. Histological glymphatic markers included aquaporin-4 (AQP4), glial fibrillary acidic protein (GFAP), podoplanin (PDPN), perivascular space (PVS) enlargement, and vessel density. High resolution MRI was used to assess a global PVS score.

**Results:** Compared with TLE-controls, TLE-ENC specimens showed increased white matter AQP4 expression and AQP4/GFAP ratio, whereas the AQP4/GFAP ratio was reduced in grey matter. PDPN expression was significantly elevated in both grey and white matter in TLE-ENC cases. MRI demonstrated greater supratentorial PVS enlargement in in ENC patients. Radiological features suggestive of IIH were identified in 46.1% of TLE-ENC patients. Compared with controls, TLE-ENC patients had shorter disease duration and lacked association with previous febrile seizures. Surgical treatment achieved seizure freedom in 70% of ENC patients at a median follow-up of 32 months.

**Interpretation:** This study provides the first characterization of glymphatic alterations in TLE-ENC-related epilepsy. Dysregulation of AQP4 and PDPN together with increased PVS burden suggests a distinct glymphatic dysfunction pattern in TLE-ENC, supporting a potential pathophysiological link among ENC formation, IIH, and epileptogenesis mediated by altered cerebrospinal fluid dynamics.

## Introduction

Temporal lobe encephaloceles (ENC) are herniations of brain tissue through defects in the skull base, most commonly involving the floor of the middle cranial fossa and the anteroinferior temporal lobe. These lesions are increasingly recognized as a potentially underdiagnosed cause of focal epilepsy, particularly in adults with drug-resistant temporal lobe epilepsy (TLE) and otherwise normal MRI findings.^1–3^ The clinical presentation typically mirrors temporal lobe seizure semiology, including autonomic, psychic, and abdominal auras, and may be associated with a later age of onset and absence of febrile seizures.^1,4,5^ Neuroimaging, especially high-resolution MRI and CT of the skull base, is essential for diagnosis, as temporal ENC can be subtle and easily missed on standard imaging protocols.^3,6–7^ EEG findings usually lateralize and localize to the temporal lobe, with seizure onset often adjacent to the ENC area.^1,5^ Surgical resection, either tailored lesionectomy or anterior temporal lobectomy in temporal lobe epilepsies associated with ENC (TLE-ENC), offers a high likelihood of seizure freedom, with outcomes comparable to standard epilepsy surgery for other lesional etiologies.^1,2,5,8,9^ Early identification and targeted surgical intervention are critical for optimal management of epilepsy associated with TLE-ENCs. Although numerous studies have explored and compared surgical techniques for TLE-ENC, a consensus on the best approach is still lacking.^1^

Moreover, the etiology of TLE-ENC is still elusive: a series of multifactorial genetic and environmental contributions are thought to be involved, especially in the acquired ones. Nonetheless, most TLE-ENC cases remain “idiopathic”.^10–12^ Radiological signs of idiopathic intracranial hypertension (IIH) are frequently associated with ENC^1,4^ and chronic or intermittent elevations in intracranial pressure, for example exacerbated by untreated obstructive sleep apnea (OSA) or intracranial hypertension due to hydrocephalus, can contribute to skull base thinning and to the development of spontaneous ENCs, with obesity serving as a possible risk factor for some of these conditions.^13–19^

Interestingly, recent works supported the evidence of glymphatic impairment in IIH through increased visible PVSs and congestion of glymphatic flow, especially in the frontal-temporal regions, which may contribute to altered cerebrospinal fluid dynamics and increased intracranial pressure.^20–22,23^ In addition, recent neuroimaging studies demonstrate that patients with TLE, including those with hippocampal sclerosis, exhibit bilateral glymphatic dysfunction compared to healthy controls, as measured by the diffusion tensor imaging-along the PVSs index.^24,25^ This dysfunction is associated with disease duration and cognitive impairment, suggesting that glymphatic impairment is a feature of epilepsy independent of structural lesions such as ENC.^24–26^

Thus, the possibility of studying the glymphatic system in patients with TLE-ENC with respect to TLE-controls appears relevant because its dysfunction has been implicated in IIH and epilepsy. PVSs have been proposed as a component of the glymphatic pathway and may become enlarged across several conditions linked to glymphatic dysfunction, including cerebral small-vessel disease, dementia, and across multiple epilepsy subtypes, further supporting the role of glymphatic system alterations in epilepsy pathophysiology.^27^ Aquaporin-4 (AQP4) is the most abundantly expressed water channel protein in the central nervous system. Localized on astrocytic endfeet, where it facilitates the exchange between cerebrospinal fluid and interstitial fluid within the brain parenchyma, AQP4 is considered an essential component of the glymphatic system.^28^ Similarly, podoplanin (PDPN), a transmembrane glycoprotein expressed in lymphatic endothelial cells has emerged as a critical marker of PVS and their associated lymphatic structures.^29^

Based on these premises, we investigated glymphatic function in TLE-ENC compared with TLE of other aetiologies. In both cohorts, we evaluated radiological and histological signs of PVS enlargement, as well as immunohistochemical expression of AQP4 and PDPN, and of GFAP and CD34, to evaluate gliosis and vascular density.

## Materials and methods

### Patients’ selection

Twenty of the 46 patients with TLE-ENC diagnosed at the IRCCS Foundation “Carlo Besta” Neurological Institute in Milan between 2009 and 2025 underwent epilepsy surgery. Thirteen of the 20 patients were enrolled according to the following inclusion criteria: *(a)* TLE; *(b)* intraoperative and/or imaging findings unequivocally consistent with a temporal lobe ENC; *(c)* anatomo-electro-clinical correlation consistent with the localization of the epileptogenic zone within or adjacent to the temporal ENC, *(d)* absence of other epileptogenic lesion including hippocampal sclerosis, *(e)* tissue available for research purpose. The control group was composed of 12 consecutive TLE patients (TLE-control) treated between 2016 and 2023, with the following inclusion criteria: *(a)* TLE; *(b)* brain MRI acquired during the pre-surgical work-up that excluded the presence of temporal ENC; *(c)* seizure freedom after epilepsy surgery as confirmation of the hypothesized epileptogenic zone; *(d)* at least two years of post-surgical follow-up; *(e)* absence of radiological signs of IIH (observed in only one excluded patient in the consecutive series) and *(f)* postsurgical tissue available for research purpose. Written informed consent was obtained for the surgical procedure and to use brain surgical specimens and clinical data for research purposes, in compliance with institutional and national ethical guidelines; the protocol for the study was approved by the local ethics committee (EpiBesta protocol n.83, 14-04-2021).

### Histologic evaluation of the postsurgical specimens

Serial 7 µm-thick paraffin-embedded sections were stained with hematoxylin and eosin, thionin, and luxol fast blue. For each case, diagnostic immunohistochemistry (IHC) was performed in accordance with the ILAE recommendation for the neuropathological workup of epilepsy surgery specimens.^30^ For the specific purpose of the present work, one representative neocortical section showing the most evident histological features of ENC and a comparable section in the TLE-control group, were additionally stained with the following antibodies: polyclonal AQP4 (Sigma, Merk Life Science, MI, Italy, diluted 1:200), monoclonal PDPN (clone D2-40; Dako, Carpinteria, CA, diluted 1:20) and monoclonal CD34 (clone QBEnq-10, Dako, diluted 1:50). For each antibody, all samples from both the TLE-ENC and TLE-control cohorts were processed in a single staining run to ensure inter-specimen consistency. IHC was performed using an avidin-biotin peroxidase method, with 3,3-diaminobenzidine (Sigma, St Louis, MO) as the chromogen, followed by hematoxylin counterstaining.

AQP4-, GFAP-, PDPN- and CD34-stained sections were digitized at 20x magnification using Aperio slide scanner (CS2 Leica Biosystem, Vista, CA, USA). Scanning was performed on the same day, in batches of five slides, using a single calibration performed at the beginning of the session to maintain identical parameters for all samples. Quantitative image analysis was carried out using Image-Pro Premier software (version 9.3; Media Cybernetics, Silver Spring, MD, USA). For each marker, two 0.46 mm² regions of interest (ROIs) were manually selected at high magnification. One ROI included the superficial grey matter (GM) in areas showing histological signs of ENC, or in anatomically comparable TLE-control regions; the second ROI was placed in the deep white matter (WM). Following hematoxylin subtraction, images were converted into binary masks (black: positively stained area; white: unstained area). Quantitative estimates of AQP4, GFAP, and PDPN immunoreactivity were obtained by applying threshold-based segmentation and by measuring the percentage of stained area within each ROI (field fraction). For CD34, the number of immunolabeled vessels was manually counted. PDPN-immunostained sections were further analyzed to assess PVS in WM vessels using QuPath software (version 0.6.0; https://qupath.github.io). ROIs encompassing the entire WM were manually delineated, and vessels with clearly identifiable endothelium were selected and classified into two categories: vessels with enlarged PVS defined by the presence of a prominent cavity between the brain parenchyma and the endothelium, and vessels with normal PVS. The percentage of vessels exhibiting enlarged PVS and the area of the perivascular cavities were subsequently calculated.

### Imaging protocol

High resolution T2-weighted images were acquired on a 3T MR scanner, with a slice thickness of 3 or 4 mm. MRI review was performed according to prior experience.^20^ PVS were assessed using a comprehensive qualitative rating scale with good inter-observer agreement, as validated by Potter *et al*. ^31^ Specifically, the radiologist scored PVS in the basal ganglia, centrum semiovale, and midbrain. Basal ganglia and centrum semiovale PVSs were graded as 0 (none), 1 (1–10), 2 (11–20), 3 (21–40), or 4 (>40) and called supratentorial PVS. Whereas midbrain PVS were rated as 0 (not visible) or 1 (visible). Finally, the sum of the two values was calculated as global PVS. Quantification was performed on T2-weighted images, while T1-weighted and FLAIR images (when available) to distinguish PVS from other lesions.

MRI studies were independently reviewed by two board-certified radiologists with 9 (F.M.D.) and 16 (V.C.) years of experience in brain imaging in epilepsy surgery cases. Discrepancies were resolved by consensus after a joint re-assessment. IIH signs were defined according to established MRI criteria and included: (1) empty or partially empty *sella turcica*, (2) distension of optic nerve sheath, (3) enlarged Meckel caves, and (4) transverse venous sinus stenosis when imaging was available. Patients were classified as having radiological signs of IIH if they exhibited at least two of the above features, consistent with prior literature.^32^

### Statistical analysis

Continuous and ordinal variables are expressed as median and interquartile range (IQR) and compared between TLE-ENC and TLE-control using the Mann-Whitney U test. Categorical variables are expressed as counts and percentages. Contingency table analysis was used to evaluate the associations between categorical variables, with the independence of the rows and columns being tested by means of Fisher’s exact test. All analyses were performed using SPSS software (IBM Corp. Released 2021. IBM SPSS Statistically for Macintosh, Version 28.0. Armonk, NY: IBM Corp). Statistical significance was set at p<0.05.

## Results

### Clinical findings

Of the 13 TLE-ENC patients included in the study, 8 were females (61.5%) and 5 were males (38.5%). The 12 TLE-control group included 5 females (41.7%). A complete summary of clinical data is shown in Table 1. No patient exhibited clinical symptoms of IIH. Five patients with TLE-ENC were treated with a standard antero-mesial temporal lobectomy and dura mater repair; five underwent cortectomy and dural reconstruction; two patients received cortectomy and tailored amygdala resection; and one patient was treated with cortectomy in the first place and later with lobectomy due to seizure recurrence. Surgical indication and the choice of approach were established in a multidisciplinary meeting based exclusively on therapeutic grounds considering anatomo-electro-clinical correlation. Regardless of the surgical approach, among patients with a minimum follow-up of two years (n=10; median 32 months, IQR 24-46), seven (70%) achieved Engel class I (five in class Ia), two class II, and one class III.

**Table 1.** Clinical features of our cohorts.

|  | TLE<sup>c</sup>-control patients | TLE-ENC<sup>d</sup> patients | p |
| --- | --- | --- | --- |
| Number of patients | 12 | 13 |  |
| Sex, female, N<sup>a</sup> (%) | 5 (41.7) | 8 (61.5) | p>0.05 |
| Handedness, right, N<sup>a</sup> (%) | 10 (83.3) | 13 (92.9) | p>0.05 |
| Age at epilepsy onset, years, median (IQR<sup>b</sup>) | 11 (2-29) | 25 (9-40) | p>0.05 |
| Age at surgery/current age, years, median (IQR<sup>b</sup>) | 44 (35-48) | 45 (32-51) | p>0.05 |
| Epilepsy duration, years, median (IQR<sup>b</sup>)* | 24 (14-37) | 11 (7-15) | p<0.01 |
| Seizure frequency, N/month, median (IQR<sup>b</sup>) | 6 (4-10) | 2 (1-5) | p>0.05 |
| Neurological examination, altered, N of patients (%) | 2 (16.7) | 1 (7.1) | p>0.05 |
| History of febrile convulsions, N of patients (%)* | 9 (75.0) + 1 doubtful | 0 | p<0.01 |
| History of dystocic delivery, N of patients (%) | 2 (16.7) | 0 | p>0.05 |
| History of perinatal distress, N of patients (%) | 3 (25.0) + 1 doubtful | 3 doubtful (23.1) | p>0.05 |
| History of head trauma, N of patients (%) | 1 (8.3) | 2 (15.4) | p>0.05 |
| Other prior events, N of patients (%) | 0 | 0 | p>0.05 |
| BMI, N, median (IQR<sup>b</sup>) | 24.16 (21.28-25.56) | 24.10 (20.25-26.45) | p>0.05 |
| Overweight, N of patients (%) | 5 (41.7) | 6 (46.2) | p>0.05 |

\*statistically significant difference among the two groups (p<0.01). All other features were comparable among the two groups.
<sup>a</sup>N: patients' number,
<sup>b</sup>IQR: interquartile range,
<sup>c</sup>TLE: temporal lobe epilepsy,
<sup>d</sup>TLE-ENC: Temporal Lobe Epilepsy associated with encephalocle

The 12 TLE-control patients were treated with standard antero-mesial temporal lobectomy and remained seizure-free after surgery with at least two years of follow-up, as per inclusion criteria. Analysis of clinical characteristics revealed significant differences between the two TLE groups regarding epilepsy history: duration of epilepsy was notably longer in TLE-control compared to TLE-ENC patients (p=0.01) and febrile seizures were documented in 9 out of 12 control patients (75%), and in none of the 13 TLE-ENC patients (p<0.01). Furthermore, in patients with TLE-ENC a tendency toward a later age of onset and lower seizures’ frequency was observed.

We found no significant differences in TLE-ENC group in clinical findings according to seizures outcome.

### Histological assessment

The 13 patients with TLE-ENC showed variable histopathological alterations, including superficial protrusion of brain parenchyma covered by meningeal cells, focal distortion of cortical architecture and gliosis.^4^ In the TLE-control group, all cases but one exhibited hippocampal sclerosis, either isolated (n=6) or associated with FCDIIIa (n=5); aspecific gliosis was found in one patient.

AQP4 immunoreactivity showed in both groups a broad distribution in the neuropil in GM and WM, with a more intense signal around blood vessels, consistent with its polarized localization at distal and perivascular astroglial endfeet. Gliosis was diffuse and more prominent in the WM. PDPN expression was mainly localized at the pial border around WM vessels, and diffusely within the WM neuropil, showing variable intensity that was particularly evident in the TLE-ENC cases. In areas where histological signs of ENC were clearly identifiable, such as brain protrusions or glial septa, the immunoreactivities were more prominent. Vascular endothelial cells were visualized using the CD34 marker, which labeled small capillaries in the GM and predominantly small arterioles in the WM. Enlargement of PVS in WM vessels was observed more frequently in the TLE-ENC group (Figs. 1, 2).

**Figure 1.**
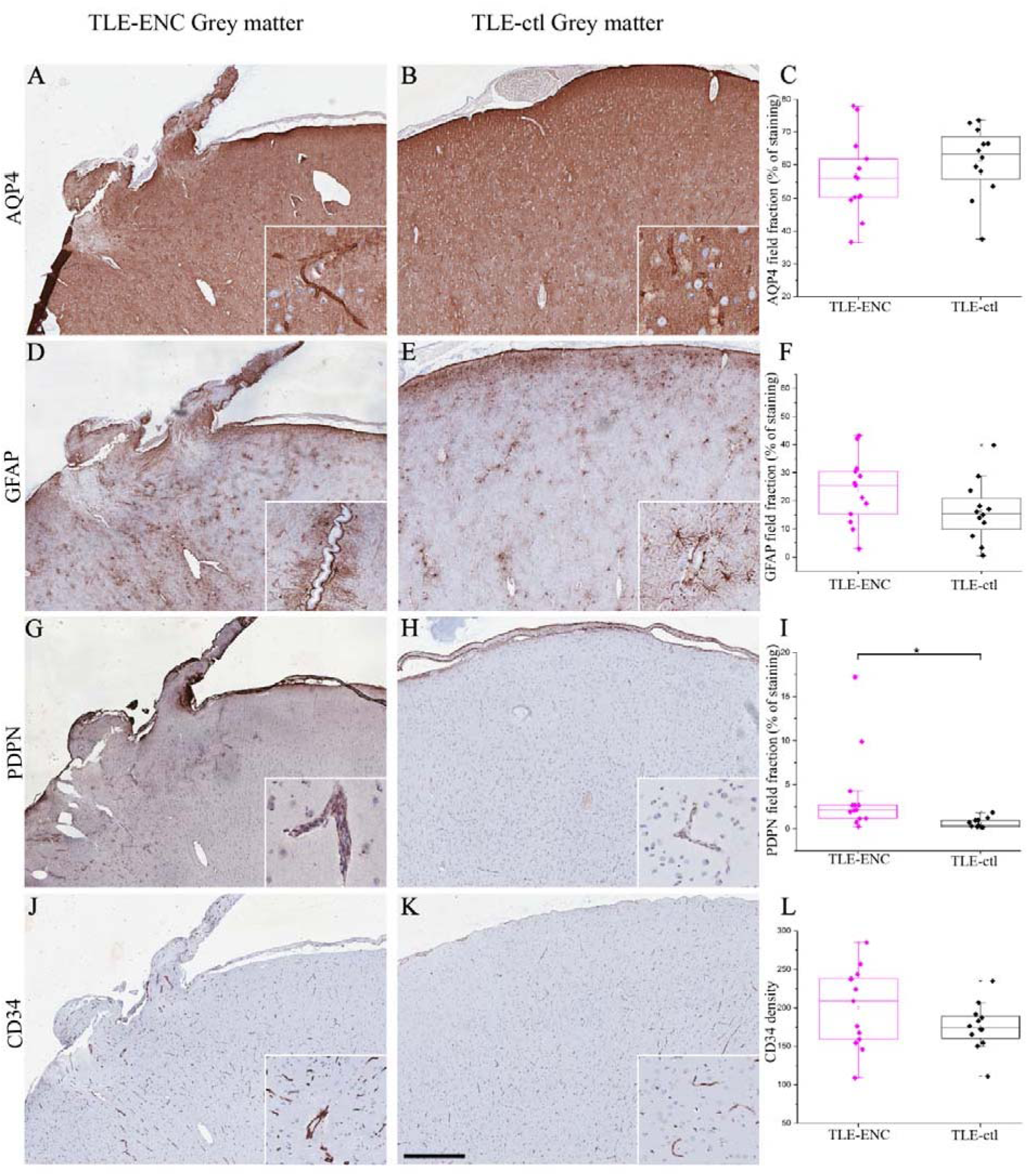
Histological markers evaluation in grey matter of TLE-ENC compared with TLE-ctl. Representative images show protrusions of the brain parenchyma in TLEt ENC tissue samples (A, D, G, J). Immunoreactivity (ir) for AQP4 (A, B), GFAP (D, E), PDPN (G, H), and CD34 (J, K) is shown for both groups, together with the corresponding quantitative analyses (C, F, I, L). Statistical significance was set at pt<t0.05. AQP4 and GFAP ir display a diffuse distribution within the neuropil and are particularly concentrated around blood vessels (insets). PDPN is predominantly localized at the pial border, while CD34 clearly highlights vascular density. Scale bar: 180 µm (A-K); 73 µm (insets).

**Figure 2.**
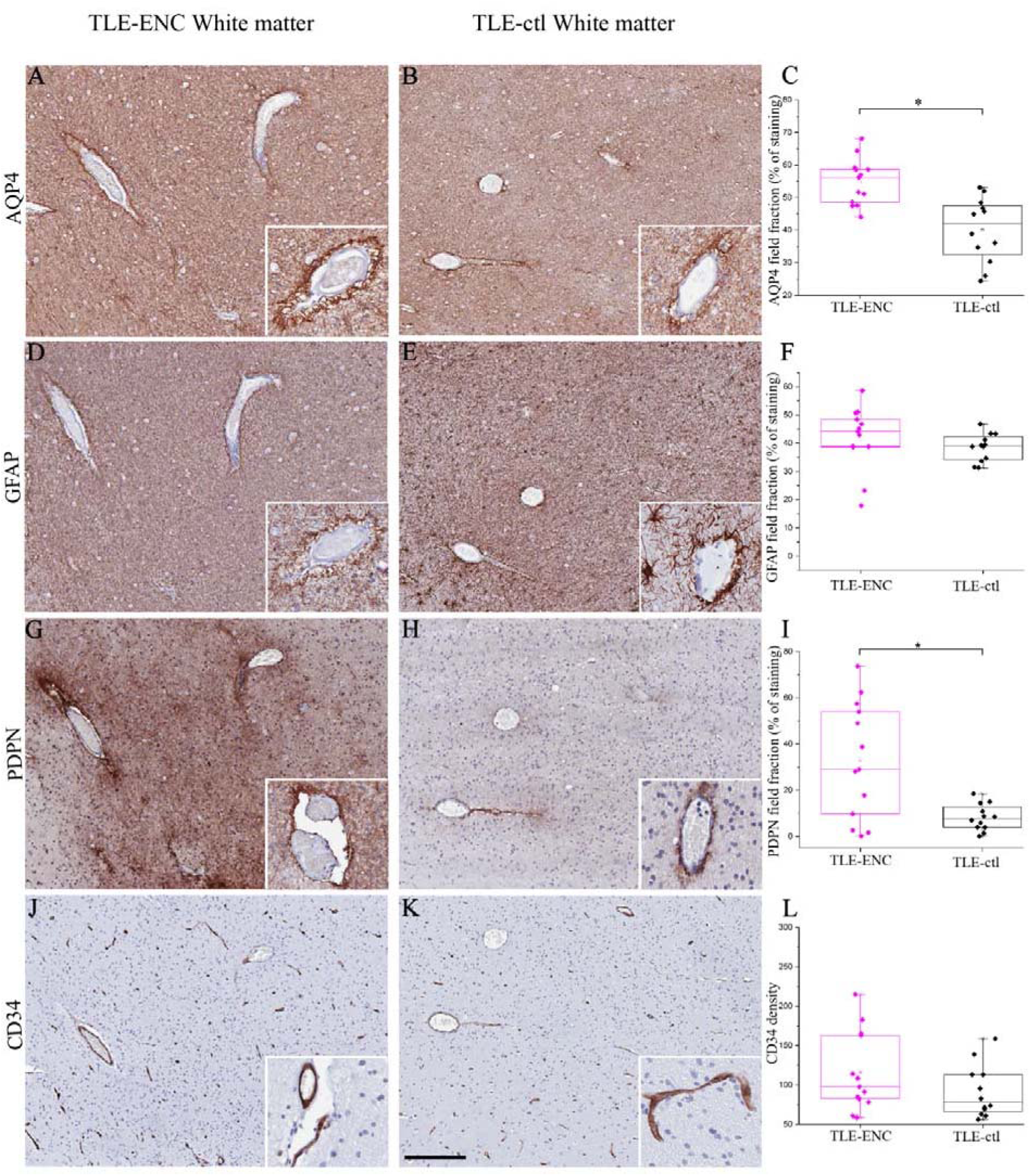
Histological markers evaluation in white matter of TLE-ENC compared with TLE-ctl. Immunoreactivity (ir) for AQP4 (A, B), GFAP (D, E), PDPN (G, H), and CD34 (J, K) is shown for both groups, together with the corresponding quantitative analyses (C, F, I, L). Statistical significance was set at pt<t0.05. AQP4, GFAP, and PDPN ir are markedly increased in the neuropil of TLEt ENC compared with TLEtctl. Blood vessels are labeled by all markers and enlargement of perivascular spaces (PVSs) is more frequently observed in the TLEtENC group (insets). Scale bar: 180 µm (A-K); 73 µm (insets).

Quantitative analysis of all immunohistochemical markers revealed no differences according to seizure outcome, therefore all analysis was performed combining all TLE-ENC cases. We found significant differences between TLE-ENC and TLE-control tissue samples (Figs. 1, 2). AQP4 expression in the WM was significantly higher in TLE-ENC specimens (median 56.1; IQR 48.6–58.6) compared with TLE-control (median 41.8; IQR 32.5–47.6; p<0.01). After correction for gliosis (AQP4/GFAP ratio), AQP4 labelling remained significantly increased in the WM of TLE-ENC samples, and showed reduced expression in the GM. Moreover, PDPN expression was significantly higher in TLE-ENC cases compared with TLE-control, both in the WM (median 28.9; IQR 9.7–53.9 *vs*. median 7.7; IQR 3.9–12.6; p<0.01) and in the GM (median 2.1; IQR 1.2–2.7 *vs*. median 0.3; IQR 0.2–0.9; p=0.02). Other immunohistochemical measurements showed trends that did not reach statistical significance: in both groups, CD34 vessel density was higher in GM that in WM, and the proportion of WM vessels exhibiting enlarged PVSs was greater in TLE-ENC compared with TLE-control. No significant differences were observed between the two TLE groups in the area of the perivascular cavities.

When the TLE-ENC group was stratified by the presence (n=6) versus the absence (n=7) of radiological IIH signs, no significant associations emerged, and surgical outcomes did not correlate with specific AQP4 and PDPN immunohistochemistry findings.

We found no significant association between duration of epilepsy and quantitative histological markers.

### Radiological results

Five out of 13 TLE-ENC patients had bilateral temporal ENCs, while eight had unilateral TLE-ENC. Six out of 13 patients (46.2%) showed MRI signs of IIH. TLE-control patients had mesial lesions, and none had IIH as per inclusion criteria. We found no significant differences in TLE-ENC group in radiological results according to seizures outcome, therefore all analysis was performed combining all TLE-ENC cases. Evaluation of PVS enlargement in supratentorial regions revealed a significant difference between the two groups (Fig. 3). TLE-control subjects exhibited a median PVS enlargement score of 2 (IQR 1-2.5), while TLE-ENC patients showed a median score of 3 (IQR 2-3), (p=0.05). TLE-ENC patients had higher global PVS enlargement scores compared to TLE-controls, though this difference approached but did not reach statistical significance (median 4, IQR 3-5 *vs*. median 3, IQR 2-4; p=0.07). When analyzing the TLE-ENC group according to presence or absence of IIH signs, no difference was found.

**Figure 3.**
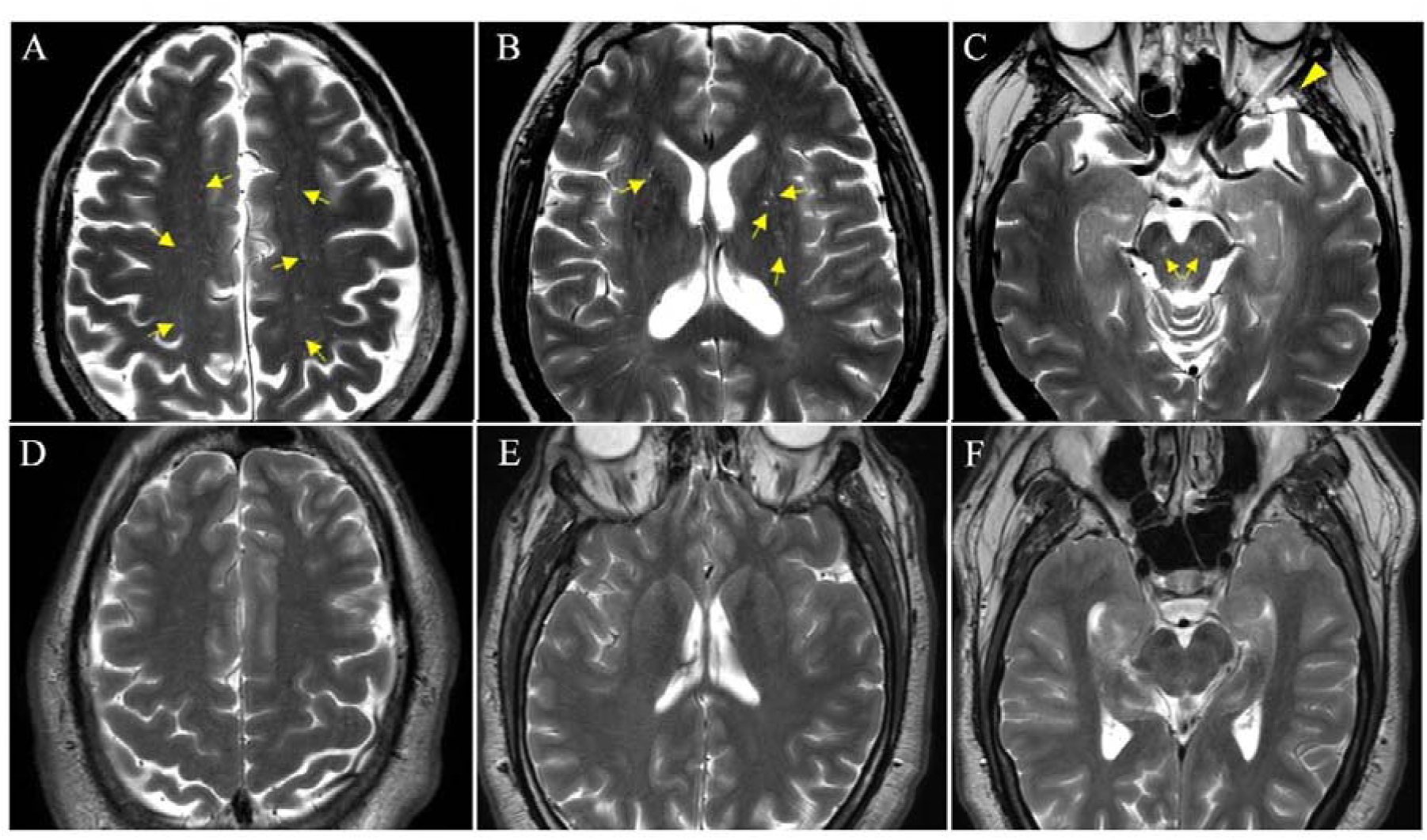
Brain MRI assessment of PVS in the basal ganglia and centrum semiovale (supratentorial regions), and in the midbrain. Top row showed axial T2-weighted images at the level of the centrum semiovale (A), basal ganglia (B), and midbrain (C) for TLE-ENC patient no.2. The left sphenoidal encephalocele is also noted (arrowhead in panel C). Bottom row showed same regions for TLE-control patient no.9 (D-E-F respectively). Multiple small dilated PVSs are observed bilaterally in the centrum semiovale, as well as a few PVSs in the basal ganglia and midbrain in the TLE-ENC patient (yellow arrows), which are absent in the TLE-control subject (only some PVSs in the centrum semiovale are highlighted).

## Discussion

The present study describes the neuropathological and imaging features of the glymphatic system in postsurgical tissue obtained from a cohort of patients with temporal ENC-related epilepsy, and compares these findings with the tissue of a control group of TLE patients with different aetiologies that achieved seizure freedom after surgery. To the best of our knowledge, this is the first study that systematically investigates glymphatic markers, including AQP4 and PDPN expression, as well as PVS alterations, in patients with TLE-ENC.

### Clinical findings

Our clinical findings are consistent with previous reports in the literature. Patients with TLE-ENC had no history of febrile seizures and exhibited a shorter duration of epilepsy compared with TLE-controls. Indeed, although it did not reach statistical significance, late-onset epilepsy was more frequently observed in the TLE-ENC than in TLE-control population.^1,2,4,33^

Among TLE-ENC patients with adequate follow-up, 70% achieved Engel class I outcomes following epilepsy surgery, consistent with previous reports.^2,8,33^ A recent systematic review reported one-year post-operative outcomes of Engel I (or ILAE 1) in 75% of patients undergoing lesionectomy, 80% after anterior temporal lobectomy with amygdalohippocampectomy, and 85% after anterior temporal lobectomy.^1^ These results underscore the importance of recognizing TLE-ENC as a surgically remediable cause of drug-resistant epilepsy. Also, as for now, there is no clear superiority of one of the above mentioned surgical approaches, and the extent of resection must be tailored to each patient based on several diagnostic findings.

### Histology

Following its high expression on the astrocytic endfeet surrounding PVSs, AQP4 is widely regarded as a reliable histological marker of the glymphatic system.^28^ This role is further supported by animal studies showing that genetic deletion of AQP4 leads to impaired glymphatic function.^34^

In human epilepsies, several alterations in AQP4 expression and localization have been reported, particularly in sclerotic hippocampi from patients with TLE.^35^ These studies demonstrated significantly increased AQP4 mRNA and protein expression compared with adjacent neocortical tissue,^35^ as well as AQP4 mislocalization characterized by loss of perivascular AQP4 and redistribution toward the neuropil.^36^ Similar alterations have also been described in human focal cortical dysplasia type IIb, a highly epileptogenic cortical lesion.^37^ Such subcellular mislocalization of AQP4 is thought to substantially affect homeostatic regulation of the neuronal microenvironment and, consequently, to contribute to neuronal hyperexcitability.^36^

In TLE-ENC cases, we observed a significantly increased expression of AQP4 in WM compared with TLE-controls, irrespective of the degree of gliosis. This finding supports the hypothesis that the observed changes reflect specific alterations in water channel expression rather than nonspecific reactive gliosis. Given that vascular density was comparable between TLE-ENC and TLE-controls within the same analyzed regions (GM or WM), and that no appreciable differences were detected in perivascular labeling, it is reasonable to speculate that the overall increase in AQP4 may be attributable to upregulation at the level of the neuropil, suggesting a possible mislocalization of AQP4 expression in ENC cases. However, the apparent preservation of perivascular AQP4 immunoreactivity should be interpreted with caution, considering the intrinsic sensitivity limitations of immunohistochemical techniques, and warrants further investigation using complementary methodological approaches.

Similarly, PDPN immunoreactivity was significantly upregulated in TLE-ENC cases in both GM and WM compared with TLE-controls. As a marker of lymphatic endothelial cells, PDPN immunoreactivity in the human brain parenchyma has been described in the PVS as part of the glymphatic system.^29^ In agreement with these observations, we detected PDPN expression in both groups at regions that are critical for lymphatic drainage, such as the pial border and around WM vessels. Diffuse neuropil immunoreactivity was observed in TLE-ENC cases, both at the histological lesions attributable to TLE-ENC and within the WM, likely reflecting a reactive astrocytic response. Consistent with this interpretation, studies in mouse models have demonstrated PDPN upregulation in reactive astrocytes following needle injury, ischemia, and tumor-associated gliosis,^38^ suggesting that this finding may be part of the functional dysregulation which disrupt their homemostatic functions.^39^

The presence of enlarged PVSs was histologically evaluated using PDPN-immunoreacted sections. However, despite a trend of increased PVS enlargement in TLE-ENC cases, histopathological findings did not reach statistical significance.

### Radiology

While histological analysis is limited by the size and location of the sample, MRI provides a broader assessment of PVS. In our analysis, MRI revealed significantly greater PVS enlargement in supratentorial regions (basal ganglia and centrum semiovale) in TLE-ENC patients compared to controls. Our findings are consistent with recent evidence from Sinclair et al.^27^ who demonstrated enlarged PVS across multiple epilepsy subtypes, with a 101%-140% increase in basal ganglia PVS volume compared to healthy controls and found no differences regarding white matter PVS. The authors proposed that enlargement of PVS in the basal ganglia may constitute a common pathophysiological feature of epilepsy, potentially reflecting either a shared mechanism underlying seizure generation or a consequence of recurrent seizures.^27^ The elevation in global PVS scores in our TLE-ENC group may underline the presence of widespread PVS alterations in ENC pathology, extending beyond regional differences. ENC-related epilepsy may thus involve systemic glymphatic impairment rather than purely focal pathology. This could be reflected in a more widespread disease expression, in line with neurophysiological observations of pathological EEG activity (mainly low-voltage fast activity) in the bilateral fronto-temporal regions and not only in the lobe where the ENC was located.^4^ The frontotemporal regions are also the sites of significant alterations in glymphatic function found with MRI study of gadobutrol clearance in patients with IIH.^23^ However, it is interesting to note that we found no clear correlation between the histological and radiological parameters of glymphatic dysfunction and the presence of radiological signs of IIH in the TLE-ENC group. This could be due to the solely qualitative dichotomous assessment (presence/absence) of signs of IIH, which may be an insensitive assessment criterion. Nonetheless, the association between TLE-ENC and IIH signs is well-established in literature.^1,33,40–42^

### Mechanistic links and therapeutic implications

The coexistence of ENC, glymphatic dysfunction, and IIH suggests a multifaceted relationship in which intracranial pressure fluctuations may represent a key, albeit non-clear, mechanistic link.^40,41^ Chronic or intermittent intracranial pressure elevations may simultaneously promote skull base remodeling, leading to ENC formation, and impair perivascular dynamics manifesting as PVS enlargement and AQP4 dysregulation. This shared pathophysiology could explain why conditions that elevate intracranial pressure, including obesity, obstructive sleep apnea, and venous outflow obstruction, are risk factors for both IIH and therefore secondary TLE-ENCs. Obstructive sleep apnea can cause transient increases in intracranial pressure through nocturnal hypoxia-hypercapnia and venous-mechanical effects, and untreated sleep apnea is associated with a higher risk of IIH and related complications, including spontaneous CSF leaks and ENCs.^43^ Similarly, OSA is associated with impaired glymphatic drainage, likely due to intermittent hypoxia and sleep fragmentation. Since the glymphatic system is most active during deep sleep and is sensitive to changes in vascular pulsatility and respiratory patterns, both OSA and IIH can disrupt its function, potentially compromising neural homeostasis. Dysfunction of the glymphatic system has been increasingly implicated in the pathophysiology of hydrocephalus.^44–46^ In hydrocephalic conditions, AQP4 expression is typically upregulated, likely as a compensatory mechanism to maintain water homeostasis in the setting of disrupted CSF circulation, though this response appears variable across different hydrocephalus subtypes.^44–48^

The present study evaluated a glymphatic signature never previously reported in ENC patients. Studying glymphatic function in TLE-ENC patients could clarify whether glymphatic dysfunction is a shared pathophysiological substrate underlying the observed association among TLE-ENC, IIH, and various risk factors of intracranial hypertension (Fig. 4). Finally, understanding this association may have important therapeutic implications, as emerging evidence demonstrates that pharmacological modulation of glymphatic fluid transport can influence disease outcomes in various neurological conditions.^49^ Specifically, pan-adrenergic inhibition has been shown to enhance glymphatic flow and suppress epileptogenesis in experimental models, while reducing reactive gliosis and preserving AQP4 polarization at astrocytic endfeet.^49^ Other promising therapeutic strategies include AQP4 facilitators, sleep optimization, and interventions targeting vascular pulsatility and meningeal lymphatic drainage.^50^ These findings suggest that targeting glymphatic dysfunction may represent a novel therapeutic avenue for patients with ENC-related epilepsy, particularly in those with concurrent IIH or sleep-disordered breathing.

**Figure 4.**
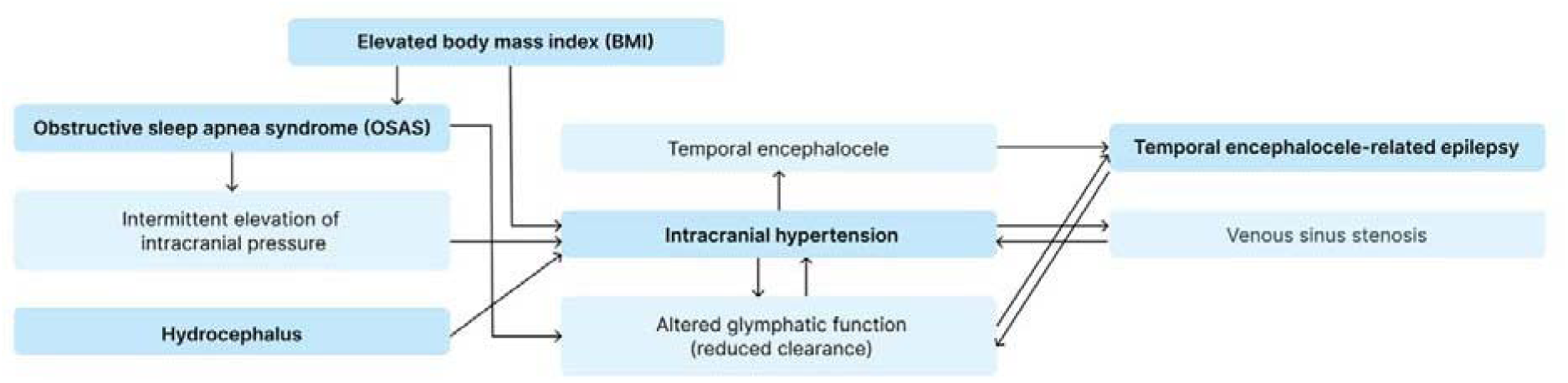
Possible links between glymphatic system, encephalocele, intracranial hypertension and its risk factors. Schematic representation of the proposed pathophysiological relationships linking elevated body mass index, obstructive sleep apnea syndrome, hydrocephalus, and venous sinus stenosis to intracranial hypertension. Increased intracranial pressure and impaired glymphatic clearance may contribute to the development of temporal encephaloceles, ultimately leading to temporal encephalocele–related epilepsy. The diagram highlights the multifactorial and bidirectional interactions among these conditions.

### Limitations

We acknowledge several limitations of the present study. The retrospective design and small sample size limit the generalizability of our findings. The control group consisted predominantly of patients with hippocampal sclerosis (11/12), which itself is associated with AQP4 dysregulation, potentially underestimating the differences between TLE-ENC and non-epileptic tissue.^35,51,52^ Systematic evaluations of OSA were not available for the entire cohort, precluding analysis of this potentially important factor. Furthermore, the qualitative assessment of IIH radiological signs may have limited sensitivity for detecting subtle associations with histological and radiological glymphatic parameters. Whether IIH represents a cause, consequence, or parallel manifestation of the same underlying glymphatic impairment remains unclear. Prospective studies comparing TLE-ENC patients with and without IIH, and IIH patients with and without ENC, are needed to disentangle these relationships. Finally, these data do not fully explain why, even in the presence of bilateral encephaloceles, seizures typically originate from only one temporal lobe. In our study, the analysis was restricted to a cohort of patients who underwent surgery for epilepsy. Consequently, our histological findings reflect the glymphatic status only within the ipsilateral hemisphere (the side of seizure onset) and may not be generalizable to the broader population of individuals with asymptomatic or non-surgical temporal encephaloceles. Further research is needed to elucidate the specific factors, whether related to degree of tethering, associated cortical dysplasia, local network effects, or other mechanisms that determine which ENC becomes the seizure focus.

### Conclusions

This study provides novel evidence of glymphatic system alterations in TLE-ENC-related epilepsy, characterized by different AQP-4 and PDPN expression and enlarged PVSs compared to TLE-controls, predominantly with hippocampal sclerosis.

Further attention to these cases is needed to better understand the etiology and pathological features of ENC-related epilepsies and to guarantee the best standard of care for patients. Clarifying the link between TLE-ENCs and glymphatic dysfunction could inform both diagnostic and therapeutic strategies, such as targeting sleep quality or CSF dynamics to improve outcomes in this patient population.

## Data Availability

All data produced in the present study are available upon reasonable request to the authors

## Data availability

The data that support the findings of this study are available from the corresponding author, upon reasonable request.

## Acknowledgments

We are grateful to the patient and their families for their participation. We thank Associazione Paolo Zorzi for supporting AB and GSC with EPICARE 2024-2026 grant. GSC was also funded by a generous donation from the Ravasio family.

